# Study on the Situation of Growth and Development of Kindergarten Children and the Relationship between Vitamin D and Body Composition, Chengdu, Sichuan Province, China

**DOI:** 10.1101/2024.07.31.24311266

**Authors:** Yanli Wang, Ying Ren, Fenglin Xing, Dan Liu, Mengjuan Hu, Luo Tongyong

## Abstract

Vitamin D plays a critical role in bone health, mineral metabolism, and overall well-being, including immune function and muscle strength. Its potential impact on body composition in children, particularly in relation to obesity and undernutrition, is an area of growing interest. However, existing literature presents inconsistent findings regarding the correlation between Vitamin D levels and body composition parameters in pediatric populations. The aim of this study was to explore the relationship between Vitamin D levels and body composition among kindergarten-aged children in Chengdu, Sichuan Province, China, providing insights that could contribute to health promotion strategies for this demographic.A cross-sectional study was conducted among children aged 3 to 6 years across 47 kindergartens in Chengdu. Participants were selected via stratified random sampling to ensure representativeness. Data collection took place during April to June to minimize seasonal effects on Vitamin D levels. Body composition was assessed using bio-electrical impedance analysis (BIA). Vitamin D status was determined through blood tests, and the distribution of Vitamin D levels by sex and age groups was analyzed. The study included 1239 children, with 75.54% having insufficient Vitamin D levels and 24.46% having sufficient levels. There was no statistically significant difference in Vitamin D status between sexes (χ^2^=0.676, p=0.411) or age groups (χ^2^=4.700, p=0.095). Correlation analysis revealed that BMI had a weak negative correlation with Vitamin D (−0.028, p=0.320) but no significant correlation with Vitamin A (−0.007, p=0.799). Similar results were observed for other body composition components. This study suggests that a majority of kindergarten children in Chengdu have insufficient Vitamin D levels, regardless of sex or age. While some correlations between Vitamin D levels and body composition were observed, they were weak and not clinically significant. Further research is needed to elucidate the complex interplay between Vitamin D, body composition, and other influencing factors in this age group. Our findings contribute to the ongoing debate about the role of Vitamin D in pediatric health and may guide future interventions aimed at improving Vitamin D status and overall body composition in children.

## 1. Introduction

Vitamin D is essential for bone health and mineral metabolism, with its roles extending into various aspects of human health, including immune function, muscle strength, and the prevention of chronic diseases. Recent research has highlighted its potential influence on body composition, including body fat percentage, muscle mass, and overall growth in children ^[1-5].^ These attributes of Vitamin D suggest that it could be a pivotal factor in addressing pediatric health issues, such as obesity and under-nutrition, which have become significant public health concerns globally.

Despite these insights, the literature presents mixed findings on the relationship between Vitamin D levels and body composition in children. Some studies have indicated that sufficient Vitamin D levels in childhood are associated with a lower body fat percentage and higher muscle mass, potentially contributing to healthier bone growth and a reduced risk of developing obesity^[6-8]^. Conversely, other studies have not found significant associations, suggesting that the relationship might be influenced by various factors including genetic predispositions, dietary intake, and physical activity levels^[9-10]^.

The current study aims to contribute to this ongoing debate by exploring the relationship between Vitamin D levels and body composition among kindergarten children in Chengdu, Sichuan Province, China. By focusing on a demographically specific population, this study seeks to provide insights that could inform both global and local health promotion strategies for children.

## 2. Materials and Methods

A cross-sectional study design was employed in this study. The retrospective data were collected from the archived physical examination of of kindergartens in Jinniu, Chengdu, Sichuan Province, China, targeting kindergarten children aged 3 to 6 years old. The research aimed to investigate the relationship between Vitamin D levels and body composition among this population.

The study received the ethics approval from the ethics committee of Jinniu maternity and child health hospital of Chengdu (2023NT01). Authors cannot access to information that could identify individual participants during or after data collection.

The kindergartens varied in size, socio-economic status of the attending families, and urban or suburban locations. Data collection occurred in three months, from April to June, to minimize the seasonal effect on Vitamin D levels.

Height and weight were measured using standardized procedures. Body composition was assessed using a bio-electrical impedance analysis (BIA) machine. Measurements of Body Mass Index(BMI), Total Body Water (TBW), Intracellular Water (ICW), Extracellular Water (ECW), Protein, Minerals, Soft Lean Mass (SLM), SLM Index (SLMI), Fat Free Mass (FFM), SMM (Skeletal Muscle Mass), PBF (Percent Body Fat), Visceral Fat Area (VFA), Body Cell Mass (BCM), Fat Free Mass Index (FFMI), Fat Mass Index(FMI), Measured Circumference of Abdomen (MCA), Skeletal Muscle Index (SMI), ECW/TBW, TBW/FFM were listed and analyzed.

Blood samples were collected by trained medical professionals to measure serum Vitamin 25-hydroxyvitamin D and Vitamin A concentrations, using a electrochemiluminescence method. Vitamin D and A assessing standards based on serum concentrations listed as below. Vitamin A Concentration: Deficiency < 0.70 μmol/L, Marginal deficiency 0.70∼1. 05 μmol/L, Sufficiency≥1.05 μmol/L. Vitamin D Concentration: Deficiency, < 30nmol/L, Insufficiency 30∼50nmol/ L, Sufficiency ≥50nmol/L ^[11].^

Children’s height, weight, and BMI, adjusted for age and sex, were categorized into eight hierarchies based on the Chinese Growth Standard for Children under 7 Years of Age (WS/T 423-2022), namely ⩾P97, ⩾P90, ⩾P75, ⩾P50, ⩾P25, ⩾P10, ⩾P3, and <P3. A height ⩾P3 indicates stunted growth, while <P3 indicates severe stunting; a weight ⩾ P3 indicates underweight, while <P3 indicates severe underweight; a BMI ⩾P97 indicates severe obesity, P90 ⩽ BMI < P97 indicates obesity, P75 ⩽ BMI < P90 indicates overweight, P3 ⩽ BMI < P10 indicates thinness, and <P3 indicates severe thinness.

Statistical analysis was performed using SPSS version 25.0. Descriptive statistics summarized the demographic characteristics of the sample. Pearson correlation analyses were conducted to explore the relationships between Vitamin D levels and various body composition measures. Independent t-tests were used to compare differences in Vitamin D levels, body composition, height, and weight across age groups and between genders. A p-value < 0.05 was considered statistically significant. Rank sum tests are used for rank data. Hierarchy analysis of height, weight, and BMI by sex and age, gender differences were assessed using the Mann-Whitney U test, while age group (3-, 4-, 5-) differences were analyzed using the Kruskal-Wallis H test.

## 3. Results

A total of 2035 children participated, with 925 girls and 1110 boys. The age distribution was 329 children in the 3-year-old group, 675 in the 4-year-old group, and 1030 in the 5-year-old group. Vitamin D and A levels were measured in a sub-sample of 1239 children.

For height, 3.20% of children were severe stunted growth, 5.31% stunted growth. For weight, 2.26% of children were severe underweight, while 5.01% were underweight. For BMI, 2.02% were severe thinness, while 5.90% thinness, whereas 5.90% were severe obesity, 6.49% obesity, and 15.0% overweight. Boys displayed significantly higher hierarchy values for height, weight, and BMI (<0.001). In age groups distribution, the overall hierarchy values of height and weight were statistical differences (p < 0.001), however no statistical difference of the overall BMI was shown (P=0.122). (Table 1).

**Table 1.**
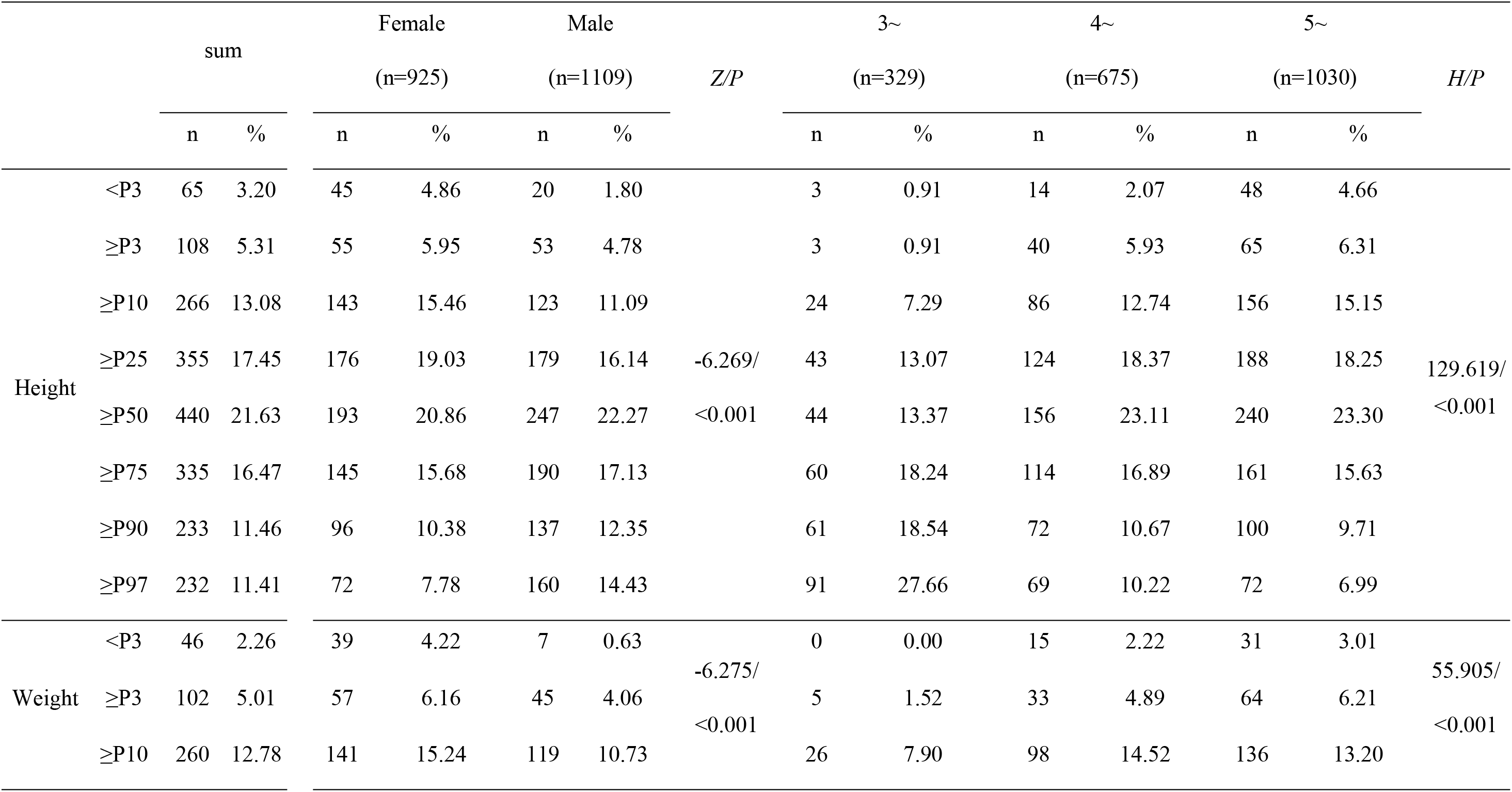
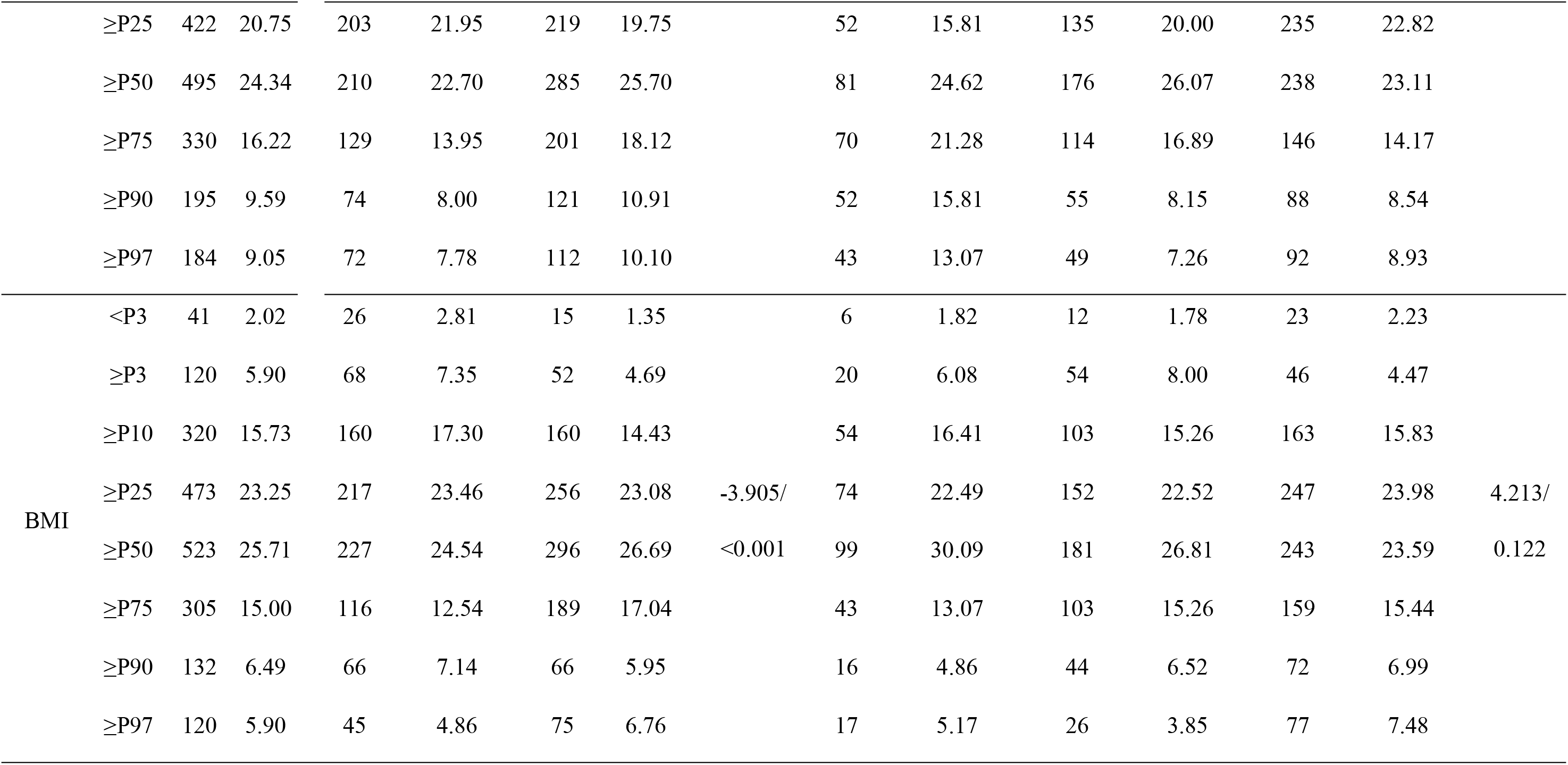
Distribution of eight hierarchies of Height, Weight and BMI by sex and age groups in 2034 children.

No significant differences in Vitamin D and Vitamin A levels were observed between boys and girls across all age groups (P=0.095 and P=0.437 respectively). Although boys exhibited a lower Vitamin D insufficiency rate (74.63%) than girls (76.65%), the difference was not statistically significant (p = 0.411). The prevalence of Vitamin A marginal deficiency was slightly higher in girls (23.35%) compared to boys (21.53%), although this was not statistically significant (p = 0.445). Vitamin D deficiency and vitamin A deficiency were not found in our study. (Table 2 and 3).

**Table 2.**
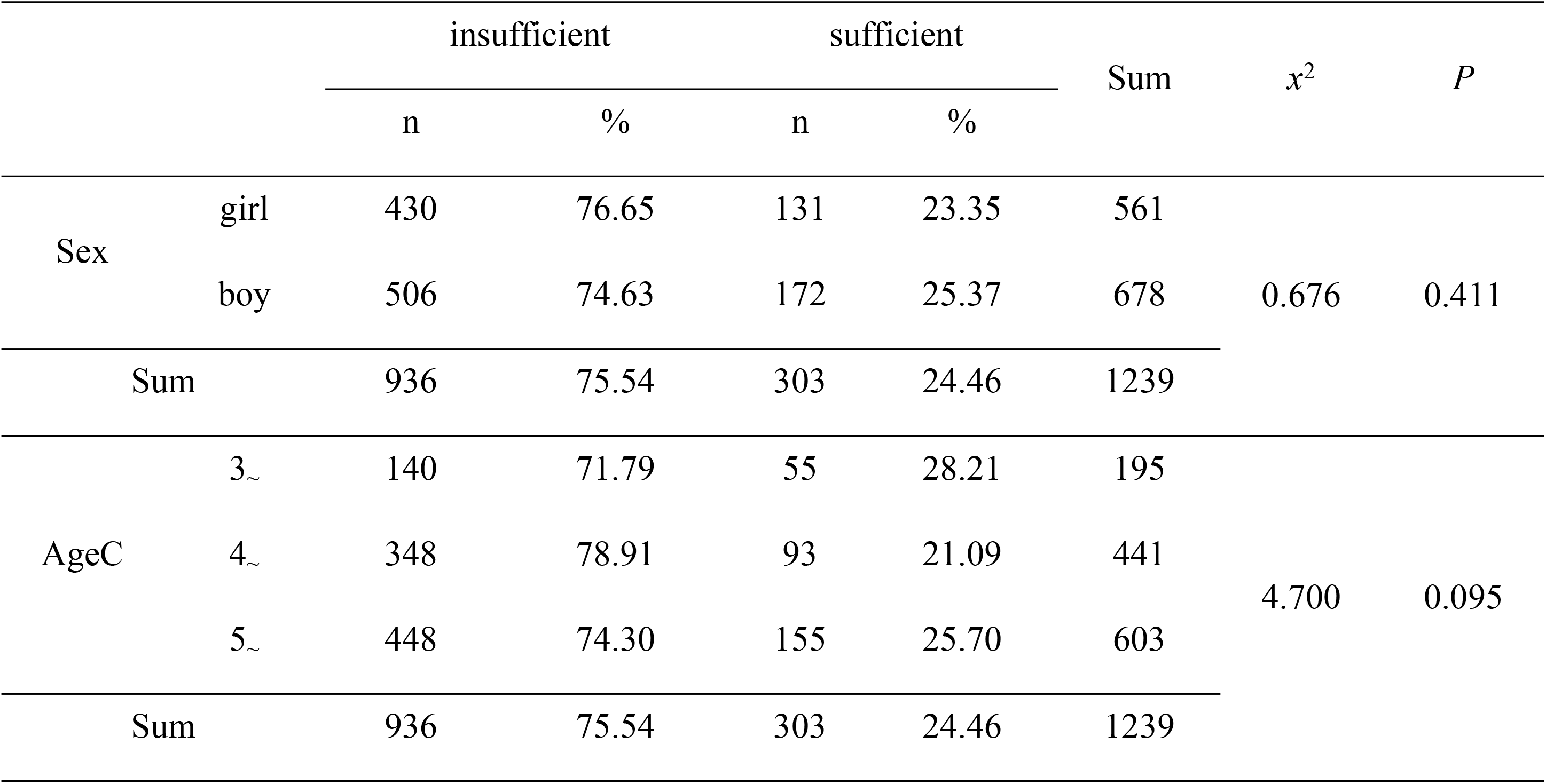
Distribution of Vitamin D status by Sex and Age groups in 1239 children.

| | | insufficient | | sufficient | | Sum | $\chi^2$ | $P$ |
| --- | --- | --- | --- | --- | --- | --- | --- | --- |
|  |  | n | % | n | % |  |  |  |
| Sex | girl | 430 | 76.65 | 131 | 23.35 | 561 | 0.676 | 0.411 |
|  | boy | 506 | 74.63 | 172 | 25.37 | 678 |  |  |
| Sum |  | 936 | 75.54 | 303 | 24.46 | 1239 |  |  |
| AgeC | 3~ | 140 | 71.79 | 55 | 28.21 | 195 | 4.700 | 0.095 |
|  | 4~ | 348 | 78.91 | 93 | 21.09 | 441 |  |  |
|  | 5~ | 448 | 74.30 | 155 | 25.70 | 603 |  |  |
| Sum |  | 936 | 75.54 | 303 | 24.46 | 1239 |  |  |

**Table 3.** Distribution of Vitamin A status by Sex and Age groups in 1239 children.

| | | marginal deficient | | sufficient | | Sum | $x^2$ | $P$ |
| --- | --- | --- | --- | --- | --- | --- | --- | --- |
|  |  | n | % | n | % |  |  |  |
| Sex | girl | 131 | 23.35 | 430 | 76.65 | 561 | 0.584 | 0.445 |
|  | boy | 146 | 21.53 | 532 | 78.47 | 678 |  |  |
| Sum |  | 277 | 22.36 | 962 | 77.64 | 1239 |  |  |
| AgeC | 3~ | 50 | 25.64 | 145 | 74.36 | 195 | 1.657 | 0.437 |
|  | 4~ | 99 | 22 | 342 | 78 | 441 |  |  |
|  | 5~ | 128 | 21.23 | 475 | 78.77 | 603 |  |  |
| Sum |  | 277 | 22.36 | 962 | 77.64 | 1239 |  |  |

Correlation between the serum Vitamin D and Vitamin A is statistically significant at the 5% significance level (t=0.038), however the level of linear association is weak (r= 0.186). (Figure 1)

**Figure 1.**
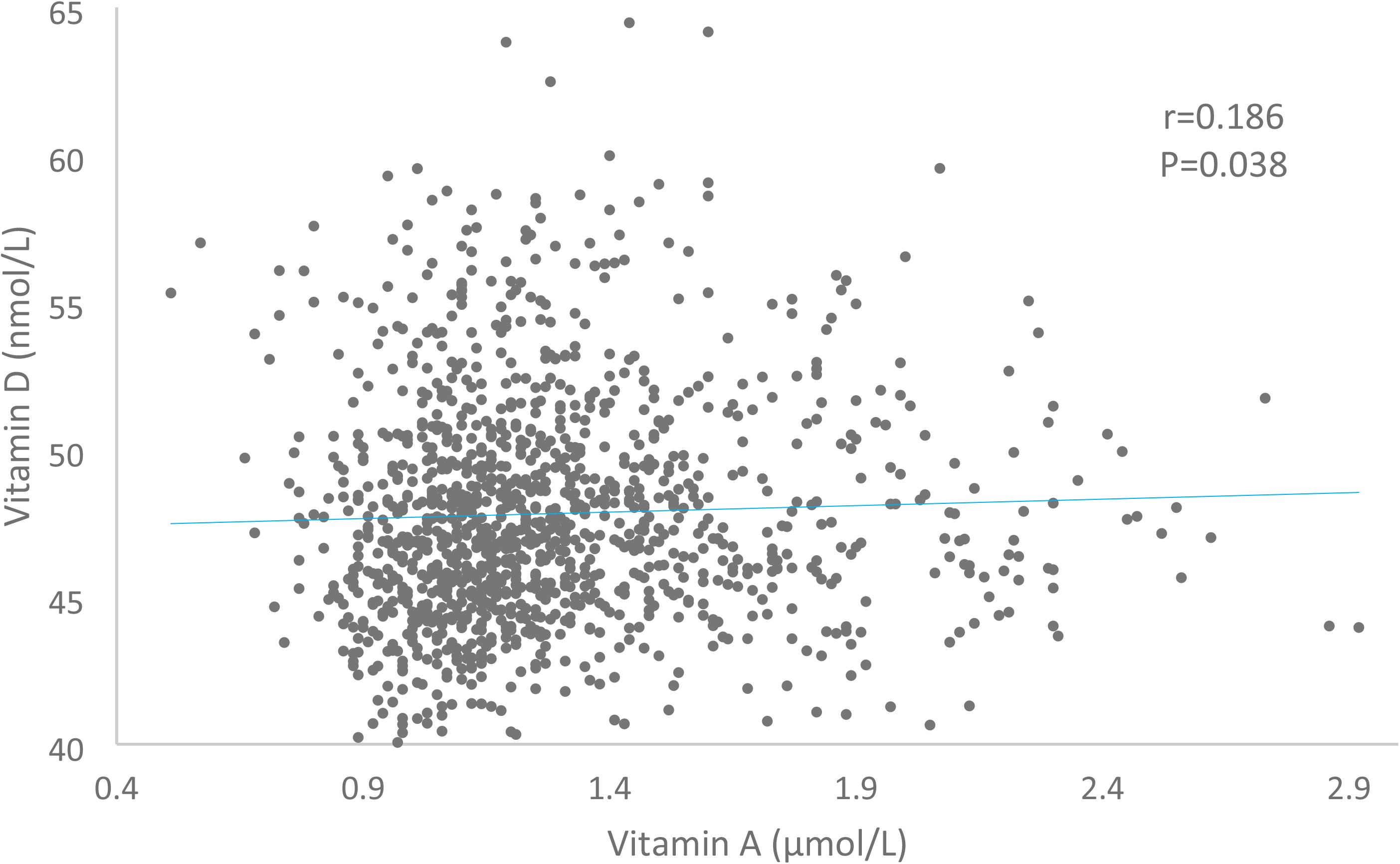
Correlation of Vitamin A and Vitamin D in 1239 children.

Except statistical association between BW/FFM, MCA and Vitamin D, there were no significant correlations between Vitamin D levels and BMI, TBW, ICW, ECW, Protein, Minerals, BFM, SLM, FFM, SMM, PBF, and other measured indices. This lack of significant association extends to Vitamin A levels across similar parameters, highlighting the complex interaction of micronutrient status with body composition in children. Table 4.

**Table 4.** Correlation analysis of body components and Vitamin D and Vitamin A in 1239 children.

|  | Vitamin D |  | VitA |  |
| --- | --- | --- | --- | --- |
|  | r | P | r | P |
| BMI (Body Mass Index) | -0.028 | 0.320 | -0.007 | 0.799 |
| TBW (Total Body Water) | 0.031 | 0.280 | -0.012 | 0.680 |
| ICW (Intracellular Water) | 0.034 | 0.232 | -0.010 | 0.718 |
| ECW (Extracellular Water) | 0.025 | 0.384 | -0.014 | 0.620 |
| Protein | 0.034 | 0.230 | -0.009 | 0.749 |
| Minerals | -0.026 | 0.357 | -0.027 | 0.342 |
| SLM (Soft Lean Mass) | 0.031 | 0.269 | -0.011 | 0.689 |
| SLM Index | 0.006 | 0.826 | -0.007 | 0.817 |
| FFM (Fat Free Mass) | 0.027 | 0.346 | -0.013 | 0.644 |
| SMM (Skeletal Muscle Mass) | 0.033 | 0.248 | -0.010 | 0.726 |
| PBF (Percent Body Fat) | -0.029 | 0.313 | 0.005 | 0.847 |
| ECW/TBW | -0.052 | 0.067 | -0.033 | 0.252 |
| VFA (Visceral Fat Area) | -0.032 | 0.255 | -0.040 | 0.155 |
| BCM (Body Cell Mass) | 0.033 | 0.242 | -0.009 | 0.747 |
| TBW/FFM* | 0.071 | 0.012 | 0.029 | 0.315 |
| FFMI (Fat Free Mass Index) | -0.019 | 0.501 | -0.008 | 0.782 |
| FMI (Fat Mass Index) | -0.032 | 0.266 | -0.001 | 0.984 |
| MCA (Measured Circumference of Abdomen)** | 0.105 | <0.001 | 0.024 | 0.391 |
| SMI (Skeletal Muscle Index) | 0.004 | 0.898 | -0.009 | 0.755 |

## 4. Discussion

The findings of this study provide a comprehensive examination of the relationship between Vitamin D levels and body composition in a significant sample of kindergarten children from Chengdu, Sichuan Province, China. We found that 75.54% of the preschoolers were Vitamin D insufficiency and 22.36% were marginal Vitamin A deficiency, but none Vitamin D deficiency and vitamin A deficiency were observed. The prevalence of Vitamin D insufficiency was lower in boys (74.63%) than in girls (76.65%), and the marginal deficiency rates of Vitamin A in boys and girls was consistent with Vitamin D (21.53% and 23.35%), but neither was statistically significant. The prevalence of vitamin D insufficiency has been reported to vary around the world, with prevalence rates of 20% in Australia, 23.3% in the United States, 34.2% in Africa, 40.4% in Europe, and 53.6% in Japan ^[12-16]^. A systematic review and meta-analysis in 2021 showed that the overall prevalence of vitamin D deficiency and insufficiency of chinese children and adolescents was 23.0% and 46.8% respectively, which means 69.8% of them were vitamin D undernourished^[17]^. It is generally consistent with the results of this study. Song P et al. reported that the prevalence of both Vitamin A deficiency and marginal Vitamin A deficiency was the highest in children aged 0-5 years at 19.53% and 28.22%, respectively, with both decreasing to 14.48% and 24.28% in children aged 6-12years in low- and middle-income countries^[18]^. The results of a systematic evaluation by the School of Public Health of Peking University in 2017 showed that the vitamin A deficiency rate among children aged 0 to 12 years in China was 5.16%, and the marginal deficiency rate was 24.29%^[19]^. This indicates that the nutritional status of vitamin A in preschool children in Chengdu is better than the national level. The prevalence estimates also declined with increasing age, from 31.53% in 0-4 years’ children to 31.68% in children aged 5-9 years, which is generally consistent with this study^[19]^.

Our research suggested gender differences in anthropometric measurements, with boys showing higher mean values for height, weight, and BMI. A cross-sectional study in Jiangsu, China showed that boys had lower rates of growth retardation, underweight and wasting than girls (1.45% vs. 1.78%, 0.54% vs. 0.67%, 1.36% vs. 2.00%) ^**[20]**^, This is consistent with our study. Wang Fuman et al. reported that the obesity rate of preschool children in seven cities in China was 12.4%^**[21]**^, which was higher than this study; and the rates of overweight and obesity in boys were higher than those in girls, with the rates increasing with age,which was consistent with the present study.This finding contributes to the growing evidence on gender disparities in growth patterns and body composition during early childhood. However, the similar rates of Vitamin D sufficiency between genders suggest that Vitamin D’s role in these differences is minimal, if existent.

Consistent with previous research^[22-24]^,our study found no significant correlation between Vitamin D levels and various measures of body composition, including BMI, BFM, SLM, and SMM. Moreover, many researchers have studied the effects of vitamin D supplementation on normal weight or overweight/obese children deficient in vitamin D, including body weight, body composition, growth and development the effects remain controversial^[25-27]^. This aligns with the body of evidence suggesting that while Vitamin D is crucial for bone health and metabolism, its impact on body fat and muscle mass in children may not be as significant as previously thought.

The lack of a significant association between Vitamin D levels and body composition in this population might be due to several factors. Firstly, genetic variations, dietary patterns, sun exposure habits, and overall health status can all influence Vitamin D metabolism and its effects on the body. Additionally, the narrow range of Vitamin D levels observed in our study population could have limited our ability to detect significant associations.

Our study found a correlation between vitamin A and vitamin D, but the correlation was weak (r=0.186, P=0.038). Multiple studies show interaction between vitamin A and vitamin D. A study in Brazilian covering 468 children suggested that serum retinol concentrations were significantly and positively correlated with vitamin D (P=0.001), and vitamin D adequacy increased up to 1.38-fold for each 1-μmol/L increase in vitamin A concentration (95% CI:1.18-1.61)^[28]^. Li XY reported that vitamin D3 only can induced catalase (CAT) activity by about 100%,when together with 9-cis retinoic acid, the CAT activity was increased by 230%^[29]^. Given the prevalence of vitamin A and vitamin D deficiencies in children and the synergistic effects of vitamin A and vitamin D, combined supplementation of the two may help to improve the nutritional status of both vitamin A and vitamin D.

Future research should aim to address these limitations by incorporating longitudinal designs, examining genetic factors that might influence Vitamin D metabolism, and considering the role of dietary Vitamin D intake and sun exposure. Understanding the complex interactions between these factors and body composition will be crucial for developing targeted interventions to improve child health outcomes.

## 5. Conclusion

This study contributes to the existing literature by exploring the relationship between Vitamin D levels and body composition among kindergarten children in Chengdu, Sichuan Province, China. While it found no significant correlation between Vitamin D levels and body composition, it highlighted important gender differences in growth patterns. These findings underscore the complexity of the relationship between Vitamin D and body composition and point to the need for further research in this area.

## Data Availability

All relevant data are within the manuscript and its Supporting Information files.

## References

1. Montenegro KR, Cruzat V, Carlessi R, Newsholme P. Mechanisms of vitamin D action in skeletal muscle. Nutr Res Rev. 2019 Dec;32(2):192–204. doi: 10.1017/S0954422419000064. Epub 2019 Jun 17. PMID: 31203824.

2. Sonne J, Kofoed-Enevoldsen P, Andersen TB, et al. Association between 25-hydroxyvitamin D level and body fat percentage in preschool children. Pediatr Obes. 2015;10(8):600–6. doi:10.1111/pedob.12225.

3. Healthier bone growth and overall growth: Aksoy N, Alagol R, Kose N, et al. The relationship between serum 25-hydroxyvitamin D levels and linear growth in healthy schoolchildren. J Int Med Res. 2011;39(5):849–54. doi:10.1177/0300926811413731.

4. Bezrati I, Hammami R, Ben Fradj MK, Martone D, Padulo J, Feki M, Chaouachi A, Kaabachi N. Association of plasma 25-hydroxyvitamin D with physical performance in physically active children. Appl Physiol Nutr Metab. 2016 Nov;41(11):1124–1128. doi: 10.1139/apnm-2016-0097. Epub 2016 Jul 13. PMID: 27764544.

5. Cediel, G., Corvalán, C., Aguirre, C. et al. Serum 25-Hydroxyvitamin D associated with indicators of body fat and insulin resistance in prepubertal chilean children. Int J Obes 40, 147–152 (2016). 10.1038/ijo.2015.148

6. Doaei S, Jarrahi S, Torki S, Haghshenas R, Jamshidi Z, Rezaei S, Moslem A, Ghorat F, Khodabakhshi A, Gholamalizadeh M. Serum vitamin D level may be associated with body weight and body composition in male adolescents; a longitudinal study. Pediatr Endocrinol Diabetes Metab. 2020;26(3):125–131. English. doi: 10.5114/pedm.2020.97466. PMID: 32901469.

7. Kouda K, Nakamura H, Fujita Y, Ohara K, Iki M. Vitamin D status and body fat measured by dual-energy X-ray absorptiometry in a general population of Japanese children. Nutrition. 2013 Oct;29(10):1204–8. doi: 10.1016/j.nut.2013.03.010. Epub 2013 Jun 22. PMID: 23800567.

8. Li H, Huang T, Xiao P, Zhao X, Liu J, Cheng H, Dong H, Morris HA, Mi J; China Child and Adolescent Cardiovascular Health (CCACH) Collaboration Group. Widespread vitamin D deficiency and its sex-specific association with adiposity in Chinese children and adolescents. Nutrition. 2020 Mar;71:110646. doi: 10.1016/j.nut.2019.110646. Epub 2019 Nov 9. PMID: 31896064

9. Kasvis P, Cohen TR, Loiselle SÈ, Hazell TJ, Vanstone CA, Weiler HA. Associations between Body Composition and Vitamin D Status in Children with Overweight and Obesity Participating in a 1-Year Lifestyle Intervention. Nutrients. 2022 Jul 30;14(15):3153. doi: 10.3390/nu14153153. PMID: 35956333; PMCID: PMC9370728.

10. White Z, White S, Dalvie T, Kruger MC, Van Zyl A, Becker P. Bone Health, Body Composition, and Vitamin D Status of Black Preadolescent Children in South Africa. Nutrients. 2019 May 31;11(6):1243. doi: 10.3390/nu11061243. PMID: 31159206; PMCID: PMC6627122

11. Chinese Preventive Medicine Association.Child Health Branch. Expert consensus on the clinical application of vitamin A and vitamin D in Chinese children (2024) [J]. Chinese Journal of Child Health, 2024, 32(4):349–358. DOI: 10.11852/zgetbjzz2024-0279.

12. Malacova E, Cheang PR, Dunlop E, Sherriff JL, Lucas RM, Daly RM, et al. Prevalence and predictors of vitamin D deficiency in a nationally representative sample of adults participating in the 2011-2013 Australian Health Survey. Br J Nutr 2019;121(8):894–904. doi: 10.1017/s0007114519000151.

13. Herrick KA, Storandt RJ, Afful J, Pfeiffer CM, Schleicher RL, Gahche JJ, et al. Vitamin D status in the United States, 2011-2014. Am J Clin Nutr 2019;110(1):150–7. doi: 10.1093/ajcn/nqz037.

14. Mogire RM, Mutua A, Kimita W, Kamau A, Bejon P, Pettifor JM, et al. Prevalence of vitamin D deficiency in Africa: a systematic review and meta-analysis. Lancet Glob Health 2020;8(1):e134–e42. doi: 10.1016/s2214-109x(19)30457-7.

15. Cashman KD, Dowling KG, Skrabakova Z, Gonzalez-Gross M, Valtuena J, De Henauw S, et al. Vitamin D deficiency in Europe: pandemic? Am J Clin Nutr 2016;103(4):1033–44. doi: 10.3945/ajcn.115.120873.

16. Nakamura K, Kitamura K, Takachi R, Saito T, Kobayashi R, Oshiki R, et al. Impact of demographic, environmental, and lifestyle factors on vitamin D sufficiency in 9084 Japanese adults. Bone 2015;74:10–7. doi: 10.1016/j.bone.2014.12.064.

17. Liu W, Hu J, Fang Y, Wang P, Lu Y, Shen N. Vitamin D status in Mainland of China: A systematic review and meta-analysis. EClinicalMedicine. 2021 Jul 14;38:101017. doi: 10.1016/j.eclinm.2021.101017. PMID: 34308318; PMCID: PMC8283334.

18. Song P, Adeloye D, Li S, Zhao D, Ye X, Pan Q, Qiu Y, Zhang R, Rudan I; Global Health Epidemiology Research Group (GHERG). The prevalence of vitamin A deficiency and its public health significance in children in low- and middle-income countries: A systematic review and modelling analysis. J Glob Health. 2023 Aug 11;13:04084. doi: 10.7189/jogh.13.04084. PMID: 37565390; PMCID: PMC10416138.

19. Song P, Wang J, Wei W, Chang X, Wang M, An L. The Prevalence of Vitamin A Deficiency in Chinese Children: A Systematic Review and Bayesian Meta-Analysis. Nutrients. 2017 Nov 25;9(12):1285. doi: 10.3390/nu9121285. PMID: 29186832; PMCID: PMC5748736.

20. Liu Y, Wang YY, Cheng Y, et al. Growth and development of children and related influencing factors: a cross-sectional study of the families with children aged 0-6 years in Jiangsu Province. Zhongguo Dang Dai Er Ke Za Zhi. 2022 Jun 15;24(6):693–698. Chinese. doi: 10.7499/j.issn.1008-8830.2202072. PMID: 35652424; PMCID: PMC9250398.

21. Wang Fuman, Yao Yi, Yang Qi. A cohort study of wasting, overweight and obesity among preschool children in seven cities in China [J]. Chinese Journal of Disease Control & Prevention, 2019, 23(5): 522–526. DOI: 10.16462/j.cnki.zhjbkz.2019.05.05.006.

22. Pruszkowska-Przybylska P, Sitek A, Rosset I, Sobalska-Kwapis M, Słomka M, Strapagiel D, Żądzińska E, Morling N. Associations between second to fourth digit ratio, cortisol, vitamin D, and body composition among Polish children. Sci Rep. 2021 Mar 29;11(1):7029. doi: 10.1038/s41598-021-86521-7. PMID: 33782473; PMCID: PMC8007693.

23. Correa-Rodríguez M, Schmidt-RioValle J, Ramírez-Vélez R, Correa-Bautista JE, González-Jiménez E, Rueda-Medina B. Influence of Calcium and Vitamin D Intakes on Body Composition in Children and Adolescents. Clin Nurs Res. 2020 May;29(4):243–248. doi: 10.1177/1054773818797878. Epub 2018 Aug 31. PMID: 30168345.

24. White Z, White S, Dalvie T, Kruger MC, Van Zyl A, Becker P. Bone Health, Body Composition, and Vitamin D Status of Black Preadolescent Children in South Africa. Nutrients. 2019 May 31;11(6):1243. doi: 10.3390/nu11061243. PMID: 31159206; PMCID: PMC6627122.

25. Ganmaa D, Bromage S, Khudyakov P, Erdenenbaatar S, Delgererekh B, Martineau AR. Influence of Vitamin D Supplementation on Growth, Body Composition, and Pubertal Development Among School-aged Children in an Area With a High Prevalence of Vitamin D Deficiency: A Randomized Clinical Trial. JAMA Pediatr. 2023 Jan 1;177(1):32–41. doi: 10.1001/jamapediatrics.2022.4581. PMID: 36441522; PMCID: PMC9706398.

26. Corsello A, Macchi M, D’Oria V, Pigazzi C, Alberti I, Treglia G, De Cosmi V, Mazzocchi A, Agostoni C, Milani GP. Effects of vitamin D supplementation in obese and overweight children and adolescents: A systematic review and meta-analysis. Pharmacol Res. 2023 Jun;192:106793. doi: 10.1016/j.phrs.2023.106793. Epub 2023 May 11. PMID: 37178775.

27. Brzeziński M, Jankowska A, Słomińska-Frączek M, Metelska P, Wiśniewski P, Socha P, Szlagatys-Sidorkiewicz A. Long-Term Effects of Vitamin D Supplementation in Obese Children During Integrated Weight-Loss Programme-A Double Blind Randomized Placebo-Controlled Trial. Nutrients. 2020 Apr 15;12(4):1093. doi: 10.3390/nu12041093. PMID: 32326621; PMCID: PMC7230345.

28. Lourenço BH, Silva LL, Fawzi WW, Cardoso MA; ENFAC Working Group. Vitamin D sufficiency in young Brazilian children: associated factors and relationship with vitamin A corrected for inflammatory status. Public Health Nutr. 2020 May;23(7):1226–1235. doi: 10.1017/S1368980019002283. Epub 2019 Aug 23. PMID: 31439064; PMCID: PMC10200414.

29. Li XY, Xiao JH, Feng X, Qin L, Voorhees JJ. Retinoid X receptor-specific ligands synergistically upregulate 1, 25-dihydroxyvitamin D3-dependent transcription in epidermal keratinocytes in vitro and in vivo. J Invest Dermatol. 1997 Apr;108(4):506–12. doi: 10.1111/1523-1747.ep12289733. PMID: 9077482.

